# Low-level mosaic variants causing the pancreatic disease congenital hyperinsulinism can be detected from blood DNA

**DOI:** 10.64898/2026.01.13.26344002

**Authors:** Jasmin J. Bennett, Thomas W. Laver, Jonna M. E. Männistö, Jayne A. L. Houghton, Elisa De Franco, Oguzhan Kalyon, Sabrina Wright, Anna-Marie Johnson, Diva D. De Leon, Evgenia Globa, Sebastian Kummer, Indraneel Banerjee, Antonia Dastamani, International Congenital Hyperinsulinism Consortium, Matthew N. Wakeling, Matthew B. Johnson, Sarah E. Flanagan

**Author notes:** Corresponding author: Prof Sarah E. Flanagan, Department of Clinical and Biomedical Science, RILD Building, University of Exeter Barrack Road, Exeter, EX2 5DW, UK. These authors contributed equally to this work (Jasmin J. Bennett, Thomas W. Laver, Matthew B. Johnson, Sarah E. Flanagan). This research was funded in whole, or in part, by Wellcome [223187/Z/21/Z]. For the purpose of open access, the author has applied a CC BY public copyright licence to any Author accepted Manuscript version arising from this submission.

## Abstract

A substantial proportion of individuals with a well-defined monogenic disorder remain without a genetic diagnosis. Low-level mosaic pathogenic variants are increasingly recognised as an underappreciated cause of monogenic disease but are technically challenging to detect, particularly in organ-specific conditions when affected tissue is inaccessible. We systematically investigated low-level mosaic variants in individuals with congenital hyperinsulinism (CHI: n=1,252) or neonatal diabetes (NDM: n=312), two opposing pancreatic disorders of insulin secretion. We screened for established pathogenic variants with variant allele fraction (VAF) <8% in dominant CHI (*ABCC8*, *GCK*, *GLUD1*, *HK1*) or dominant NDM (*ABCC8*, *KCNJ11*, *INS*) genes in targeted next generation sequencing (tNGS) data using Mutect2. This called 40 variants across the four genes in 39 individuals with CHI. No candidate variants were found in the NDM cohort. Orthogonal validation of 35 variants using TaqMan-based droplet digital PCR (ddPCR) confirmed 26/35 variants. The median VAF for confirmed variants was 3.6% (1.1–7.8%), while false positives (9/35) predominantly had a VAF <1% with some overlap in VAF with true positives. This study shows that disease-causing low-level mosaic variants in dominant CHI genes can be detected in blood using tNGS but require orthogonal validation. These results provide a framework to improve diagnostic yield in organ-specific conditions where mosaic variants may represent an important missed cause of disease.

## Introduction

A genetic diagnosis of monogenic disease provides knowledge of recurrence risk, facilitates prenatal testing and can allow personalised treatment. Despite the rapid acceleration in genetic discoveries driven by next-generation sequencing (NGS), up to 50% of individuals with a well-defined disease phenotype suggestive of a monogenic disorder lack a genetic diagnosis.^1^ Several factors may account for these genetically unresolved cases, including polygenic or phenocopy presentations, pathogenic variants in yet-to-be-identified disease genes, variants in non-coding regulatory regions that remain challenging to detect or interpret, and technical or biological limitations that hinder the identification of variants within known disease genes.

Low-level mosaic variants, typically defined by a variant allele fraction (VAF) of <10%, represent a class of disease-causing variants that are particularly challenging to detect using standard genomic approaches^2,3^ These variants arise post-zygotically and are therefore present in only a subset of a patient’s cells. In organ-specific diseases where the affected tissue is not readily accessible and analysis relies on peripheral blood sampling,^4^ detection of mosaic variants is especially challenging. For example, in the *PIK3CA* overgrowth syndrome, activating variants are often detectable only in affected tissues such as adipose, muscle, skin, or bone, whilst being absent or having a VAF too low to be captured by standard genetic testing of leukocyte DNA.^2^ Similarly, in congenital hyperinsulinism (CHI), a genetically heterogeneous disorder characterised by inappropriate insulin secretion from the pancreas leading to persistent hypoglycaemia,^5^ mosaic variants in genes such as *ABCC8, GCK* and *HK1* have been identified in pancreatic tissue from patients, but were not detected in their matched blood samples.^6–9^

Even when DNA from the affected tissue is available or the variant is detectable from leukocyte DNA, accurately identifying low-level mosaic variants remains a substantial challenge. These variants can be difficult to distinguish from background fluorescence in Sanger sequencing or from base-call errors inherent to short-read NGS. Moreover, PCR amplification, integral to both Sanger and short-read NGS library preparation, can introduce artefactual nucleotide changes that are hard to distinguish from true low-level variants. Low-level sample contamination presents another significant obstacle, further complicating reliable detection.^10^

For all monogenic disorders, and particularly those where a genetic diagnosis directly guides treatment and medical management such as in CHI and the opposing condition neonatal diabetes (NDM), distinguishing true low-level mosaic variants from technical artefacts or contamination is essential.^5,11–13^

In this study, we aimed to use targeted next-generation sequencing (tNGS) to identify causative low-level mosaic variants in whole blood DNA from a large cohort of individuals with CHI or NDM. We validated candidate low-level variants using an orthogonal method, developing a robust approach for distinguishing true pathogenic variants from technical artefacts.

## Methods

### Participants

We studied 1,252 individuals with genetically unsolved CHI and 312 individuals with genetically unsolved NDM, who had been referred to the Exeter Genomics Laboratory for genetic testing. Informed consent was obtained from each participant or their parent/guardian. Ethical approval for this study was granted by the Wales Research Ethics Committee 5 through the Genetic Beta-Cell Research Bank (22/WA/0268).

### Targeted next generation sequencing

All individuals had previously undergone routine tNGS using DNA extracted from peripheral blood leukocytes. This analysis had excluded disease-causing homozygous, heterozygous and high-level mosaic variants in the coding regions, intron/exon boundaries and relevant non-coding regions of at least 13 established disease genes for the CHI cohort, and 22 established disease genes for the NDM cohort.^14^ A list showing the minimum genes screened in both cohorts is provided in **Supplementary Table 1**.

### Calculation of variant calling threshold

Standard germline variant calling tools such as GATK HaplotypeCaller are not designed to detect low-level variants.^15^ To establish the minimum VAF threshold at which HaplotypeCaller can reliably call variants, we performed a dedicated VAF analysis on a subset of the Exeter tNGS dataset (n=10,018 samples). We compared variant calls generated by GATK HaplotypeCaller with those from GATK3 Mutect2, a tool specifically designed for detecting low-level mosaic variants.^16,17^

VAFs were calculated from VCF files using *bcftools* to extract allele depth and total depth fields, followed by custom *awk* scripts to compute the ratio of alternative to total reads at each variant site. Only variants with a total read depth ≥300 were included in the analysis. The resulting VAF distributions for both callers were explored and visualised in R using the *ggplot2* package.

### Low-level mosaic variant calling

We next screened tNGS data generated during routine genetic testing for CHI or NDM for low-level mosaic variants using GATK3 Mutect2, focusing on variants below the detection threshold of HaplotypeCaller. In all 1,252 individuals with genetically unresolved CHI, variants were called in the coding regions of the established CHI genes: *ABCC8*, *GCK* and *GLUD1.* The 46 bp minimal region of the *HK1 cis*-regulatory element was screened in 271 of these individuals.^18^ In the 312 individuals with genetically unsolved NDM, variant calling was performed across the coding regions of the established NDM genes: *ABCC8*, *KCNJ11*, and *INS*. These two sets of genes were prioritised because they follow a dominant inheritance pattern and harbour recurrent pathogenic variants. The mean read depth across all the genes/loci was ≥390X for 90% of samples (details provided in **Supplementary Table 2**).

Variant calls were filtered against a curated list of over 200 disease-relevant pathogenic or likely pathogenic variants in these genes. These variants were selected because they act dominantly and because samples from heterozygous controls were available for orthogonal testing (**Supplementary Tables 3 and 4**).

Finally, given the release of an updated version of GATK, we assessed whether this update improved Mutect2’s ability to detect low-level variants by reanalysing the dataset using the GATK4 pipeline and comparing the resulting variant calls.

### Screening of low-level mosaic CHI variants in control cohorts

To assess whether low-level variants called in the tNGS data might have arisen coincidentally, we screened tNGS data from a control cohort of 522 individuals with genetically solved CHI for low-level mosaic CHI-causing variants in *ABCC8*, *GCK*, and *GLUD1* referred to Exeter. We also screened for these same variants in the genetically unsolved NDM cohort (n=312).

### Contamination screening

As contamination by other human DNA can result in the presence of low VAF variants within a sample, we screened for contamination using VerifyBamID.^19^ The contamination threshold was set at 2%, which was >2 standard deviations above the mean contamination level across all samples.

### Droplet-digital PCR (ddPCR)

To provide orthogonal confirmation of low-level mosaic variants identified by tNGS, we used fluorescent TaqMan hydrolysis probe-based ddPCR.^20,21^ This technique partitions each reaction into up to 20,000 individual PCR droplets, enabling highly accurate quantification of VAFs. Fresh aliquots of leukocyte DNA from the same samples previously analysed by tNGS were used to validate putative low-level mosaic variants. In one case, buccal-cell DNA was also analysed. Serial dilutions of heterozygous positive control DNA into wildtype DNA (25%, 5%, and 2% VAF) were included to confirm detection sensitivity at low fractional abundances. Each sample was tested in duplicate (**Supplementary Figure 1**).

### Clinical data

The age at diagnosis of CHI and the presence/absence of hyperammonaemia was collated from standardized referral forms completed by clinicians at referral for genetic testing. This included individuals with confirmed low-level mosaic variants and individuals with heterozygous dominant CHI-causing variants in *ABCC8* (n=110), *GCK* (n=8), *GLUD1* (n=68) and *HK1* (n=58). Statistical analysis was performed in GraphPad Prism version 10. The Mann-Whitney test was used to compare the median age at diagnosis, and the Fisher’s exact test was used to compare the proportion of individuals with hyperammonaemia.

## Results

### Variants below 8% will not be reliably called by standard analysis

We found that variants with a VAF below 8% were not reliably detected by GATK HaplotypeCaller (**Figure 1**). We therefore set 8% as the upper limit to define low-level mosaic variants, as variants with allele frequencies below this threshold are likely to be missed by conventional analysis pipelines.

**Figure 1:**
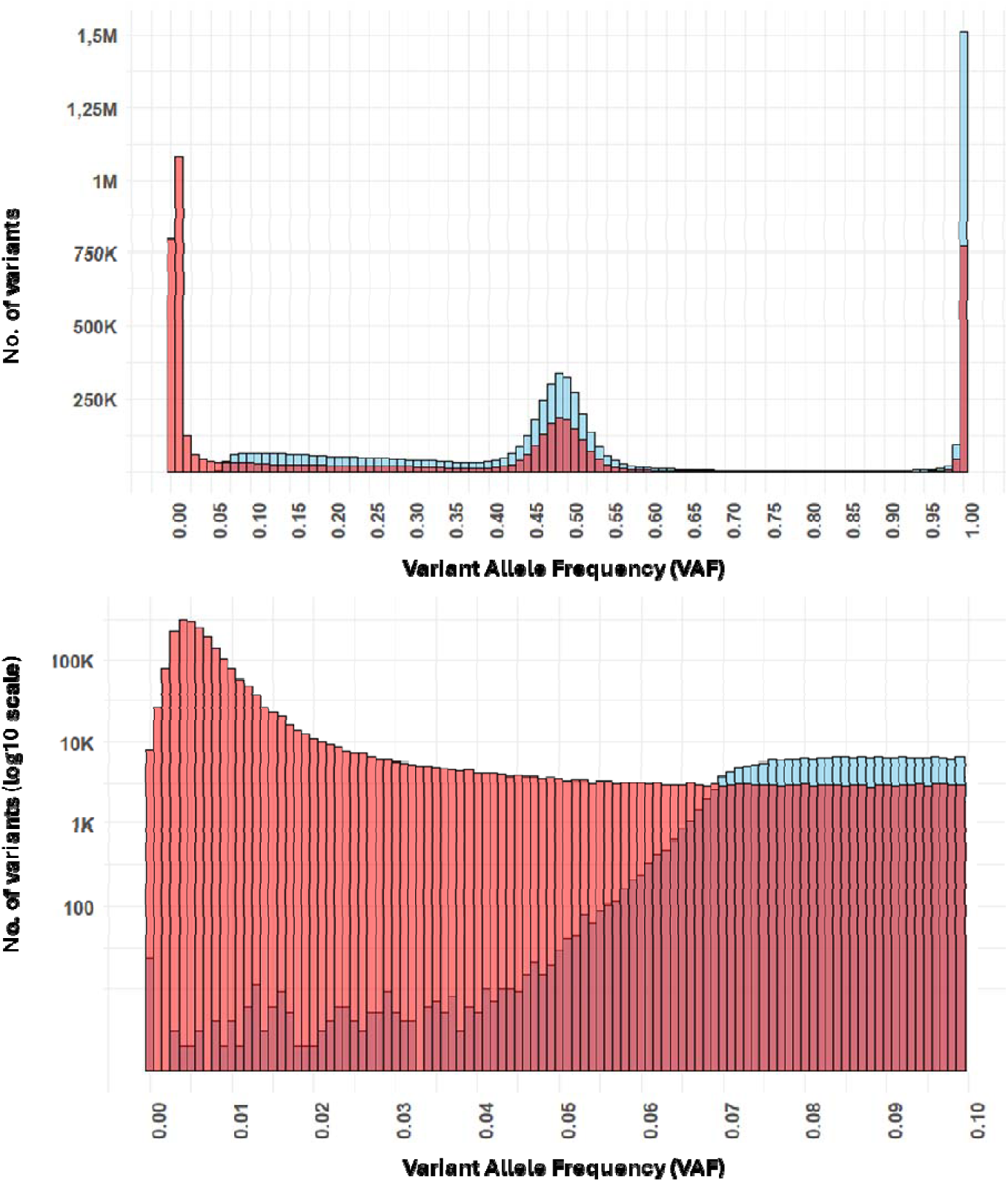
Distribution of variant allele frequencies (VAFs) from GATK haplotypecaller (blue) and Mutect2 (red) calls in targeted Next-Generation Sequencing (tNGS) data. This figure shows the distribution of VAFs for variant calls from tNGS data in 10,018 samples, where a minimum sequencing depth of 300X was achieved. The upper plot illustrates the full VAF range (0 to 100%). The lower plot provides a zoomed-in view of the VAFs between 0 and 10%, highlighting the detection limit for germline variant calling. A decrease in the number of germline calls is observed below 8%, followed by a sharp decrease below 7%.

### Low-level mosaic candidate variants called in 39 individuals from tNGS data

We identified 40 candidate low-level CHI variants (25 distinct variants) in 39 individuals with genetically unsolved CHI. The median VAF was 1.4% (range: 0.5–7.8%), supported by a median of 9 sequencing reads (range: 3–80 reads) (**Supplementary Table 5**). No candidate NDM variants were called in the genetically unsolved NDM cohort. The called CHI variants included 10 *ABCC8* variants in 12 individuals, 5 *GCK* variants in 7 individuals, 8 *GLUD1* variants in 16 individuals, and 2 *HK1* variants in 5 individuals. One individual with genetically unsolved CHI had a low-level mosaic variant called in both *ABCC8* and *HK1*.

### Detection of contamination in two samples

Contamination screening of tNGS data from the 39 individuals with a called variant revealed evidence of low-level contamination in two samples. In the first individual, with an estimated contamination level of 10.8%, an *ABCC8* variant had been called at VAF 1.1% (7/635 reads). In the second case, with an *ABCC8* variant called at 0.9% (6/682 reads), there was an estimated contamination level of 2.4%.

### 74% of candidate low-level mosaic variants confirmed by ddPCR

We performed orthogonal testing of 35 of the called low-level variants using ddPCR in individuals with genetically unsolved CHI. This analysis included the two samples where contamination had been detected. Four of the remaining variants not tested had insufficient DNA for ddPCR confirmation and the patients had been lost to follow-up. For one variant, a robust ddPCR assay could not be designed due to the sequence context (**Supplementary Table 5**).

Twenty-six of the 35 variants (74%) tested were confirmed by ddPCR (*ABCC8* n=2, *GCK* n=5, *GLUD1* n=14, *HK1* n=5) (**Figure 2, Supplementary Figure 2**, **Supplementary Table 5**). These consisted of 15 distinct variants. The median tNGS VAF of the 26 true positives was 3.6 (range: 1.1-7.8%), supported by a median of 18 sequence reads (range: 4-80 reads).

**Figure 2:**
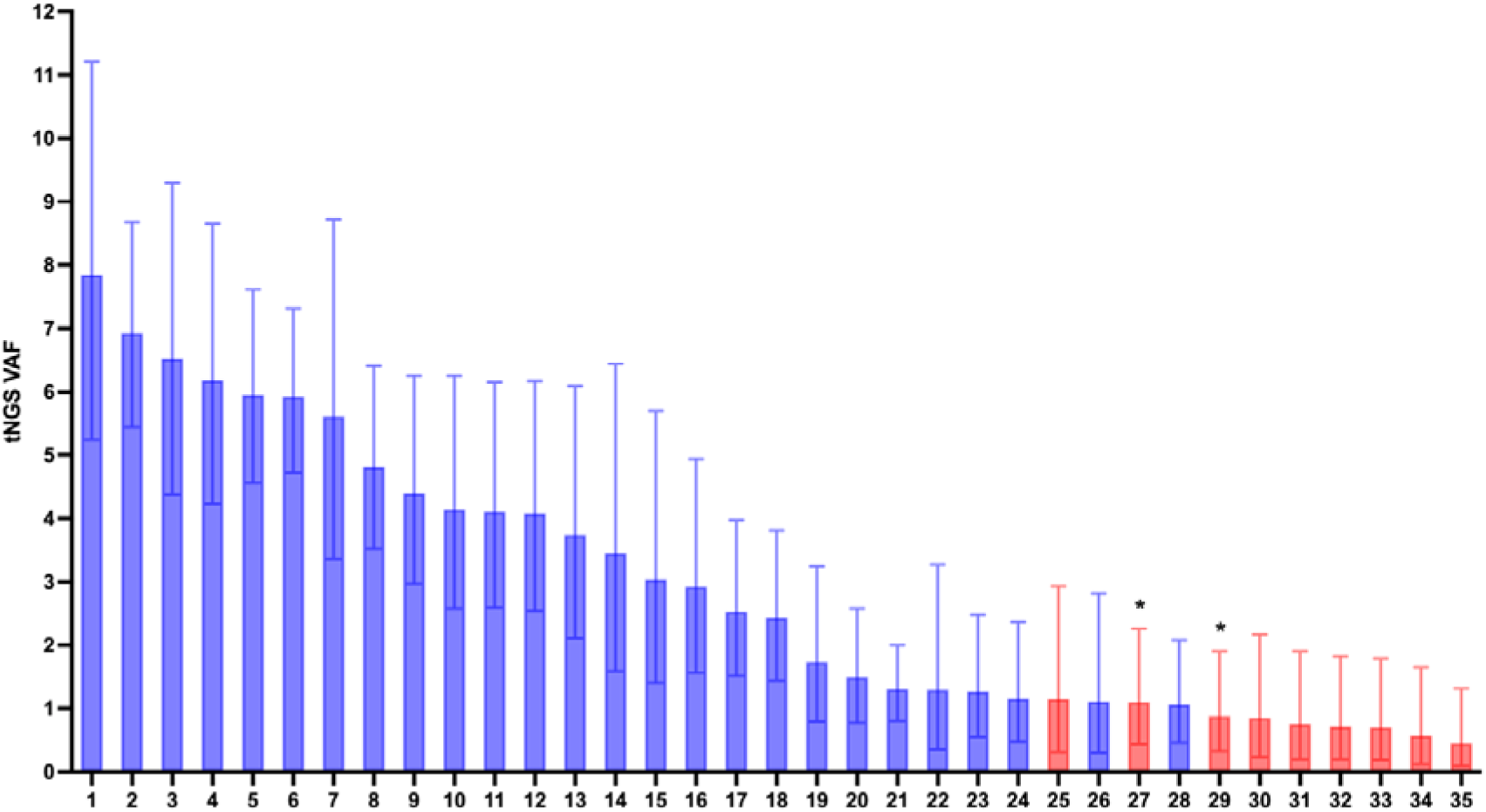
Targeted next generation sequencing data (tNGS) variant allele frequency (VAF) of the known disease-causing dominant CHI variants that were identified as candidate low-level variants in individuals with genetically undiagnosed CHI. Error bars indicate 95% confidence intervals. Blue: true positive variants confirmed by droplet digital PCR (ddPCR). Red: false positive variants not confirmed by ddPCR. * Indicates samples with evidence of contamination.

The nine variants not confirmed by ddPCR included eight variants in *ABCC8* (seven distinct variants) and one *GCK* variant (**Supplementary Table 5**). These included the *ABCC8* variant called in the individual with a *HK1* low-level variant (the *HK1* variant was confirmed by ddPCR) and the two *ABCC8* variants identified in samples that showed evidence of contamination (**Supplementary Table 5**). The median tNGS VAF for variants not confirmed was 0.8% (range: 0.5-1.2%), supported by a median of 4 reads (range: 2-7 reads).

### Mosaicism confirmed across tissues in one individual

For one individual, ddPCR showed the mosaic *HK1* variant was at similar levels in the leukocyte and buccal cell DNA, supporting mosaicism across tissues (leukocyte DNA: ∼0.7% (95% CIs: 0.5-0.9%); buccal DNA: ∼0.3% (95% CIs: 0.1-0.6%)) (**Supplementary Figure 3**).

### Low-level mosaic variants are rare in controls

A low-level mosaic *ABCC8* variant (c.4519G>A, p.(Glu1507Lys); VAF 1.2%, 2/169 reads) was called in one individual from the CHI genetically solved control cohort. This person had a previously reported heterozygous disease-causing *GLUD1* variant (c.1387A>G, p.(Asn463Asp)). No dominant CHI-causing variants in the *ABCC8*, *GCK* or *GLUD1* genes were detected in individuals with NDM. The low level mosaic CHI variants were therefore significantly enriched in individuals with genetically unsolved CHI (n=35/1,252), compared to individuals with genetically solved CHI (n=1/522) or NDM (n=0/312) (*p* <0.0005). The low-level mosaic *ABCC8* variant called in the individual with the *GLUD1* heterozygous variant was not confirmed by ddPCR.

### GATK4 Mutect2 misses more true positive low-level mosaic variants, but has higher positive predictive value than GATK3 Mutect2

To assess the ability of GAKT4 and GATK3 to accurately detect low-level mosaic variants using Mutect2, we compared the variant calls within the genetically unsolved CHI tNGS dataset. GATK4 did not identify any additional candidate low-level mosaic variants. However, GATK4 Mutect2 missed 8 of 26 (31%) true positive variants that were detected by GATK3.

Despite this, GATK4 showed a higher positive predictive value (GATK4: 90% 18/20 vs GATK3: 74% 26/35), generating fewer false positives (**Supplementary Table 5**).

### Clinical characteristics of individuals with true positive low-level mosaic variants

The median age at diagnosis of CHI appeared to be later in individuals with low-level mosaic variants in *ABCC8*, *GCK,* and *HK1* compared to those with a heterozygous dominant variant in the same gene (*ABCC8*: 21 days vs 1 day, *GCK*: 256 days vs 0 days, *HK1*: 213 days vs 7 days, **Table 1**); however these differences did not reach statistical significance (*p* >0.05). In contrast, the age at diagnosis was similar between the *GLUD1* groups (147 days for low level mosaic variants vs 183 days for heterozygous variants; **Table 1**).

**Table 1:** Comparison of clinical features between individuals with congenital hyperinsulinism (CHI) with a confirmed low-level mosaic variant versus individuals with CHI with a heterozygous dominant variant, split by gene. Continuous data are presented as median [IQR] (range), Categorical data are presented as % (n). The number of individuals with data available is indicated by n. *P* values were calculated using the Mann-Whitney statistical test for continuous data and Fisher’s exact test for categorical data.

| Clinical feature | Gene | Confirmed low-level mosaic CHI variant | Heterozygous dominant CHI variant | *P* value |
| --- | --- | --- | --- | --- |
| Age at diagnosis, days | *ABCC8* | 21 [14-28] (7-35)<br>n=2 | 1 [1-3] (0-913)<br>n=56 | 0.07 |
|  | *GCK* | 256 [139-338] (7-365)<br>n=4 | 0 [0-29] (0-4015)<br>n=18 | 0.1 |
|  | *HK1* | 213 [144-304] (84-730)<br>n=5 | 7 [0-259] (0-9490)<br>n=58 | 0.07 |
|  | *GLUD1* | 147 [77-351] (7-546)<br>n=14 | 183 [62-311] (0-2920)<br>n=68 | 0.8 |
| Hyperammonaemia |  | 54 (7/13)<br>n=13 | 92 (49/53)<br>n=53 | 0.003 |

As hyperammonaemia is a common feature of *GLUD1*-CHI, we assessed its prevalence across both groups. Among the 13 individuals with confirmed low-level mosaic *GLUD1* variants where ammonia had been measured, seven (54%) had hyperammonaemia reported at referral for genetic testing. This proportion was significantly lower than that observed in individuals with heterozygous *GLUD1* variants (92%, 49/53, *P* = 0.003) (**Table 1**).

## Discussion

We identified true low-level mosaic pathogenic dominant variants in 26 individuals with genetically unsolved CHI, with a VAF of 1.1% to 7.8%, across four different genes. These variants had remained undetected during prior routine testing of leukocyte DNA. The absence of any confirmed low-level CHI variants in our control datasets, combined with the presence of hyperammonaemia in seven individuals with low-level mosaic *GLUD1* variants, provides strong evidence that the variants are disease-causing.

The variants detected in this study are presumed to be present at low levels in leukocyte DNA rather than derived from circulating ectopic pancreatic DNA. Although the precise cellular origin of the variants was not confirmed there is evidence for their presence in multiple tissues. This includes the presence of hyperammonaemia, a kidney-associated phenotype,^22^ in seven individuals with low-level mosaic *GLUD1* variants, and the detection of a low-level *HK1* variant in buccal cell DNA of one child, which is predominantly epithelial in origin. This variant has recently been detected at a level of 1.2% in the pancreatic DNA of the same child by another team.^23^ Further evidence supporting tissue-wide mosaicism comes from a prior report describing an individual with a *GLUD1* variant detected at 2.7% VAF in leukocyte DNA whose child had inherited the pathogenic variant, confirming its presence in germ cells.^24^ Single-cell methylation analysis could further elucidate the cellular origin of the mutated DNA detected in this study.^25^

Previous studies of pancreatic tissue from children with CHI have identified variants at similarly low levels, suggesting that blood VAF may be a reasonable proxy for pancreatic mutation load.^6,7^ This is supported by comparable mosaicism levels observed in leukocyte, buccal cell, and pancreatic DNA from one individual in this study.^23^ Together, these findings indicate that over-secretion of insulin by a relatively small proportion of beta-cells is sufficient to cause clinically significant hypoglycaemia, consistent with observations in focal hyperinsulinism and insulinoma, where excessive insulin secretion from a subset of beta-cells leads to profound hypoglycaemia.^26^

This study provides some preliminary evidence that, according to the age at onset of CHI, low-level mosaic variants may generally be associated with a milder phenotype than heterozygous dominant variants in these genes. Individuals with low-level mosaic *ABCC8*, *GCK,* and *HK1,* variants showed a trend toward later CHI diagnosis, although this did not reach statistical significance, likely due to small sample size. Consistent with this, a significantly lower proportion of individuals with low-level mosaic *GLUD1* variants had hyperammonaemia compared to those with heterozygous variants, supporting a milder phenotype and aligning with previous reports.^27^ As additional cases are identified, larger cohorts will allow assessment of whether this trend persists and enable variant-specific comparisons across genotypes.

A major challenge in screening for low-level mosaic variants is distinguishing true positives from technical artefacts or sample contamination. Initial analysis of our tNGS dataset identified 40 variants across four CHI genes, nine of which were later demonstrated to be false positives. Seven of these had the lowest VAFs (<1%); however, because VAFs overlapped between true-positive and false-positives, this measure alone is insufficient to define which calls are true, highlighting the need for orthogonal validation. Two unconfirmed variants arose from tNGS data showing evidence of contamination. In one sample, the contamination level (10.8%) was inconsistent with the low VAF (1.1%), suggesting a sequencing artefact. In the second case, the VAF was approximately half the detected contamination level, so contamination could not be excluded as the cause of the low-level call. Notably, none of the ddPCR-confirmed low-level variants had tNGS VAFs below 1% (half of 2%, assuming contamination by a heterozygous carrier), indicating that stock DNA contamination is unlikely to explain their dectection.^19,28^

As laboratories transition from tNGS to whole-exome and whole-genome sequencing, detection of low-level mosaic variants in routine testing is likely to decline due to lower sequencing read-depth.^29^ Our findings also show that variant calling varies by GATK version, highlighting further challenges in balancing improved sensitivity of true positives (GATK3 Mutect2) against reduced false-positives (GATK4 Mutect2). Furthermore, although ddPCR is highly sensitive for orthogonal testing, it’s per-variant design and high labour demands limit scalability. Targeted capture approaches incorporating single-molecule molecular inversion probes or adapters containing unique molecular identifiers may provide a more scalable alternative for validating low-level mosaic variants but require further validation.^30–32^

To enrich for true positive calls, we restricted analysis to variants with established dominant effects in the most common disease-genes, with assessment further limited by the need for a heterozygous control for each ddPCR assay. Even with these constraints, nine of the 39 variants (23%) identified by tNGS in the unsolved CHI cohort were artefactual. Given the strict criteria applied, the identification of 26 variants in 1,252 individuals (2%) with genetically unsolved CHI likely represents a minimum prevalence of low-level mosaic variants in this cohort. This is particularly relevant for *HK1*, which was screened in only 271 individuals after the regulatory region was added to the panel following its recent discovery.^33^ Additional individuals may have mosaic dominant variants in the analysed genes that are below detection limits in leukocyte DNA, novel variants, or variants in other CHI genes not assessed here.

Our stringent screening and orthogonal validation gave us confidence that the identified variants were disease-causing, allowing results to be communicated to families via their clinicians. Relaxing these criteria, for example, to include all novel missense variants, would make clinical interpretation challenging, particularly given the difficulties already encountered with novel heterozygous missense variants in genes like *ABCC8*. Specifically, for the nine children with false positive calls, an incorrect genetic diagnosis could have led to inappropriate clinical management and genetic counselling regarding recurrence risk, as well as missed opportunities for further testing to identify the true underlying cause of their condition.

The absence of low-level mosaic pathogenic variants in our NDM cohort likely reflects fundamental differences in disease mechanisms and is consistent with mosaicism having previously been detected only in unaffected parents and not children with NDM.^34–36^ To cause diabetes, a much larger proportion of pancreatic beta-cells likely needs to be impaired before hyperglycaemia appears, as the remaining functional beta-cells can compensate.

This is consistent with the Eisenbarth model of type 1 diabetes and post-mortem studies of beta-cell mass in recent onset type 1 diabetes cases in which 80-90% of beta-cells are lost before hyperglycaemia becomes clinically apparent.^37,38^ These observations suggest that disorders driven by pathological overproduction or overactivation (such as CHI and PIK3CA-related overgrowth) are more sensitive to the phenotypic effects of low-level mosaicism than loss-of-function disorders. This is further supported by the presence of hyperammonaemia in individuals with low-level mosaic *GLUD1* variants, where increased glutamate dehydrogenase activity accelerates the conversion of glutamate to alpha-ketoglutarate reaction, releasing ammonia.^27^

In conclusion, low-level mosaic variants are an important cause of CHI that can be detected from routine blood-derived DNA, increasing diagnostic yield without requiring access to pancreatic tissue. This is particularly relevant for *GLUD1*-, *HK1-*, dominant *ABCC8*-, and *GCK*-CHI where most individuals respond to therapy and do not require pancreatic surgery. More broadly, our findings suggest that screening for low-level mosaicism in blood may improve diagnosis in other monogenic conditions driven by pathological overproduction or overactivity.

## Acknowledgments

SEF has a Wellcome Trust Senior Research Fellowship (Grant Number 223187/Z/21/Z). TWL is supported by the Academy of Medical Sciences/the Wellcome Trust/the Government Department of Science Innovation and Technology/the British Heart Foundation/Diabetes UK Springboard Award [SBF009\1135]. JMEM is the recipient of a European Society for Paediatric Endocrinology (ESPE) Research Fellowship and the Foundation for Paediatric Research Postdoctoral Fellowship. EDF is the recipient of a European Foundation for the Study of Diabetes/Novo Nordisk Foundation Future Leader Award. MBJ is the recipient of a Diabetes UK/Breakthrough T1D RD Lawrence fellowship (23/0006516), and this study was supported by the National Institute for Health and Care Research Exeter Biomedical Research Centre. The views expressed are those of the author(s) and not necessarily those of the NIHR or the Department of Health and Social Care. We are grateful to Congenital Hyperinsulinism International (a 501©3 organisation) who supported the costs of routine diagnostic genetic testing in Exeter for some of the individuals reported in this study through the Open Hyperinsulinism Genes Project.

## Funding

This work was supported by a research grant from the University of Pennsylvania Orphan Disease Center in partnership with the Team CHIbra and Congenital Hyperinsulinism International [MDBR-23-020-CHI]. The research was funded in whole, or in part, by Wellcome [223187/Z/21/Z]. For the purpose of open access, the author has applied a CC BY public copyright licence to any author accepted manuscript version arising from this submission.

## Data availability

The raw sequencing data generated during the current study are not publicly available to preserve patient confidentiality. Variant call format (.vcf) files are available through collaboration to experienced teams working on approved studies examining the mechanisms, cause, diagnosis and treatment of diabetes and other beta cell disorders. Requests for collaboration will be considered by a steering committee following an application to the Genetic Beta Cell Research Bank (IRAS: 316,050, https://www.diabetesgenes.org/current-research/genetic-beta-cell-research-bank/). Contact by email should be directed to Sarah Flanagan.

## Notes

### Competing Interest Statement

The authors have declared no competing interest.

### Author Declarations

Ethical approval for this study was granted by the Wales Research Ethics Committee 5 through the Genetic Beta-Cell Research Bank (22/WA/0268).

