## Supplementary Information for "Low-level mosaic variants causing the pancreatic disease congenital hyperinsulinism can be detected from blood DNA"

**Supplementary Table 1:** Minimum gene list in which disease-causing homozygous, heterozygous and high-level mosaic variants had been excluded in individuals with congenital hyperinsulinism and neonatal diabetes.

| **Disease** | **Genes screened** |
| --- | --- |
| Congenital Hyperinsulinism | *ABCC8, CACNA1D, GCK, GLUD1, HADH, HK1 (non-coding regulatory region), HNF1A, HNF4A, INSR, KCNJ11, PMM2, SLC16A1 (promoter),* and *TRMT10A* |
| Neonatal Diabetes | *KCNJ11*, *ABCC8, INS, EIF2AK3, FOXP3, GATA4, GATA6, GCK, GLIS3, HNF1B, IER3IP1, NEUROD1, NEUROG3, NKX2-2, PDX1, PTF1A, RFX6, SLC2A2, SLC19A2, STAT3, WFS1,* and *ZFP57* |

**Supplementary Table 2:** Mean read depth of targeted next generation sequencing (tNGS) data over the exons of *ABCC8*, *GCK*, *GLUD1*, *KCNJ11,* and *INS*, and the non-coding regulatory region of *HK1* in samples from individuals with genetically undiagnosed congenital hyperinsulinism (CHI, n=1,252) and neonatal diabetes (NDM, n=312).

| **Gene** | **Disease (n samples)** | **Minimum mean read depth for 90% of samples** |
| --- | --- | --- |
| *ABCC8* | CHI (n=1,252) and NDM (n=312) | 400 |
| *GCK* | CHI (n=1,252) | 400 |
| *GLUD1* | CHI (n=1,252) | 413 |
| *HK1* (non-coding 46bp region of interest) | CHI (n=271) | 581 |
| *KCNJ11* | NDM (n=312) | 278 |
| *INS* | NDM (n=312) | 394 |

**Supplementary Table 3:** Curated list of known dominant disease-causing congenital hyperinsulinism variants screened for in this study, where a heterozygous control was available allowing for ddPCR testing. The coding location of coding variants are provided according to the following transcripts *ABCC8*: NM_001287174.1, *GCK*: NM_000162.3, *GLUD1*: NM_005271.4. The genomic locations of the reported *HK1* variants in the CHI-causing region are provided according to GRCh38.

| **Gene** | **Variant** | **Evidence** |
| --- | --- | --- |
| *ABCC8* | Q474R, c.1421A>G | Reported in ^1^ |
| *ABCC8* | A478D, c.1433C>A | Reported in ^1^ |
| *ABCC8* | V715M, c.2143G>A | Reported in ^2^ |
| *ABCC8* | G716D, c.2147G>A | Reported in ^3^ |
| *ABCC8* | E825K, c.2473G>A | Reported in ^3^ |
| *ABCC8* | A1153V, c.3458C>T | Reported in ^4^ |
| *ABCC8* | A1153T, c.3457G>A | Reported in ^5^ |
| *ABCC8* | R1353H, c.4058G>A | Reported in ^6^ |
| *ABCC8* | G1384E, c.4151G>A | Reported in ^1^ |
| *ABCC8* | S1387del, c.4160_4162del | Reported in ^7^ |
| *ABCC8* | S1387F, c.4160C>T | Reported in ^3^ |
| *ABCC8* | L1390R, c.4169T>G | Reported in ^8^ |
| *ABCC8* | L1431F, c.4291C>T | Reported in ^8^ |
| *ABCC8* | Q1459E, c.4375C>G | Reported in ^8^ |
| *ABCC8* | Q1459H, c.4377G>C | Reported in ^1^ |
| *ABCC8* | G1478V, c.4433G>T | Reported in ^7^ |
| *ABCC8* | G1479A, c.4436G>C | Reported in ^9^ |
| *ABCC8* | G1479R, c.4435G>A | Reported in ^8^ |
| *ABCC8* | G1485E, c.4454G>A | Reported in ^10^ |
| *ABCC8* | G1485R, c.4453G>A | Reported in ^11^ |
| *ABCC8* | G1488R, c.4463A>G | Reported in ^4^ |
| *ABCC8* | D1506E, c.4518C>A | Reported in ^10^ |
| *ABCC8* | D1506E, c.4518C>G | Reported in ^3^ |
| *ABCC8* | D1506H, c.4516G>C | Reported in ^12^ |
| *ABCC8* | D1506N, c.4516G>A | Reported in ^4^ |
| *ABCC8* | E1507K, c.4519G>A | Reported in ^13^ |
| *ABCC8* | A1508P, c.4522G>C | Reported in ^8^ |
| *ABCC8* | I1512T, c.4535T>C | Reported in ^14^ |
| *ABCC8* | M1514K, c.4541T>A | Reported in ^10^ |
| *ABCC8* | A1537V, c.4610C>T | Reported in ^8^ |
| *ABCC8* | R1539Q, c.4616G>A | Reported in ^8^ |
| *GCK* | S64Y, c.191C>A | Reported in ^15^ |
| *GCK* | T65I, c.194C>T | Reported in ^15^ |
| *GCK* | T65A, c.193A>G | Reported in ^16^ |
| *GCK* | G68V, c.203G>T | Reported in ^17^ |
| *GCK* | S69P, c.205T>C | Reported in ^18^ |
| *GCK* | K90T, c.269A>C | Reported in ^19^ |
| *GCK* | V91L, c.271G>T | Reported in ^18^ |
| *GCK* | W99R, c.295T>A | Reported in ^15^ |
| *GCK* | T103S, c.308C>G | Reported in ^20^ |
| *GCK* | M197I, c.591G>A | Reported in ^21^ |
| *GCK* | Y214C, c.641A>G | Reported in ^22^ |
| *GCK* | V389L, c.1165G>C | Reported in ^20^ |
| *GCK* | R447L, c.1340G>T | Reported in ^23^ |
| *GCK* | A454dup, c.1361_1363dup | Reported in ^21^ |
| *GLUD1* | R274C, c.820C>T | Reported as R221C in ^24^ |
| *GLUD1* | H315Y, c.943C>T | Reported in ^25^ |
| *GLUD1* | R318K, c.953G>A | Reported as R265K in ^24^ |
| *GLUD1* | R322H, c.965G>A | Reported as R269H in ^24^ |
| *GLUD1* | N463D, c.1387A>G | Reported as N410D in ^24^ |
| *GLUD1* | N463T, c.1388A>C | Reported as N410T in ^24^ |
| *GLUD1* | P489L, c.1466C>T | Reported as P436L in ^24^ |
| *GLUD1* | Q494R, c.1481A>G | Reported as Q411R in ^26^ |
| *GLUD1* | S498L, c.1493C>T | Reported in ^27^ |
| *GLUD1* | G499S, c.1495G>A | Reported in ^27^ |
| *GLUD1* | G499R, c.1495G>C | Reported as G446R in ^26^ |
| *GLUD1* | G499A, c.1496G>C | Reported as G446A in ^28^ |
| *GLUD1* | G499V, c.1496G>T | Reported in ^29^ |
| *GLUD1* | A500T, c.1498G>A | Reported as A447T in ^26^ |
| *GLUD1* | H507Y, c.1519C>T | Reported in ^27^ |
| *HK1* | 10:69348869-69348890delins19 | Reported as 10:71108625_71108646delins19 in ^30^ |
| *HK1* | 10:69348885-69348921delins5 | Reported as 10:71108641_71108677delinsAGTAT in ^30^ |
| *HK1* | 10:69348886G>T | Reported as 10:71108642G>T in ^30^ |
| *HK1* | 10:69348889T>C | Reported as 10:71108645T>C in ^30^ |
| *HK1* | 10:69348891C>G | Reported as 10:71108647C>G in ^30^ |
| *HK1* | 10:69348891C>T | Reported as 10:71108647C>T in ^31^ |
| *HK1* | 10:69348891C>A | Reported as 10:71108647C>A in ^30^ |
| *HK1* | 10:69348892C>A | Reported as 10:71108648C>A in ^31^ |
| *HK1* | 10:69348892C>G | Reported as 10:71108648C>G in ^31^ |
| *HK1* | 10:69348892C>T | Reported as 10:71108648C>T in ^31^ |
| *HK1* | 10:69348892del | Reported as 10:71108648del in ^30^ |
| *HK1* | 10:69348895_69348912del | Reported as 10:71108651_71108668del in ^30^ |
| *HK1* | 10:69348904_69348932del | Reported as 10:71108660_71108688del in ^30^ |
| *HK1* | 10:69348908T>G | Reported as 10:71108664T>G in ^30^ |
| *HK1* | 10:69348909C>G | Reported as 10:71108665C>G in ^31^ |
| *HK1* | 10:69348909del | Reported as 10:71108665del in ^30^ |
| *HK1* | 10:69348928-69348929del | Reported as 10:71108684_71108685del in ^30^ |
| *HK1* | 10:69348931T>C | Reported as 10:71108687T>C in ^30^ |
| *HK1* | 10:69348932_69348935del | Reported as 10:71108688_71108691del in ^31^ |

**Supplementary Table 4:** Curated list of known dominant *ABCC8* and *KCNJ11* gain-of-function variants and dominant *INS* neonatal diabetes variants screened for in this study, where a heterozygous control was available allowing for ddPCR testing. The location of variants are provided according to NM_001185098.2 (*INS*), NM_001287174.1 (*ABCC8*) and NM_000525.3 (*KCNJ11*).

| **Gene** | **Variant** | **Evidence** |
| --- | --- | --- |
| *ABCC8* | I49F, c.145A>T | Reported in ^32^ |
| *ABCC8* | V86A, c.257T>C | Reported in ^33^ |
| *ABCC8* | F132L, c.394T>C | Reported in ^34^ |
| *ABCC8* | F132V, c.394T>G | Reported in ^35^ |
| *ABCC8* | L135P, c.404T>C | Reported in ^36^ |
| *ABCC8* | P206L, c.617C>T | Reported in ^4^ |
| *ABCC8* | E208K, c.622G>A | Reported in ^35^ |
| *ABCC8* | D209N, c.625G>A | Reported in ^37^ |
| *ABCC8* | D209E, c.627C>A | Reported in ^38^ |
| *ABCC8* | Q211K, c.631C>A | Reported in ^39^ |
| *ABCC8* | D212N, c.634G>A | Reported in ^38^ |
| *ABCC8* | D212Y, c.634G>T | Reported in ^33^ |
| *ABCC8* | D212G, c.635A>G | Reported in ^4^ |
| *ABCC8* | D212E, c.636C>G | Reported in ^4^ |
| *ABCC8* | V215I, c.643G>A | Reported in ^40^ |
| *ABCC8* | L225P, c.674T>C | Reported in ^35^ |
| *ABCC8* | A235V, c.704C>T | Reported in ^4^ |
| *ABCC8* | R306H, c.917G>A | Reported in ^36^ |
| *ABCC8* | V324M, c.970G>A | Reported in ^38^ |
| *ABCC8* | V360A, c.1079T>C | Reported in ^4^ |
| *ABCC8* | L451P, c.1352T>C | Reported in ^38^ |
| *ABCC8* | S532G, c.1594A>G | Reported in ^41^ |
| *ABCC8* | F536S, c.1607T>C | Reported in ^4^ |
| *ABCC8* | F577L, c.1731T>G | Reported in ^4^ |
| *ABCC8* | I585T, c.1754T>C | Reported in ^42^ |
| *ABCC8* | V587D, c.1760T>A | Reported in ^4^ |
| *ABCC8* | R826W, c.2476C>T | Reported in ^36^ |
| *ABCC8* | G833D, c.2498G>A | Reported in ^43^ |
| *ABCC8* | H863Y, c.2587C>T | Reported in ^44^ |
| *ABCC8* | H1024Y, c.3070C>T | Reported in ^45^ |
| *ABCC8* | N1123D, c.3367A>G | Reported in ^46^ |
| *ABCC8* | E1141G, c.3422A>G | Reported in ^4^ |
| *ABCC8* | A1153G, c.3458C>G | Reported in ^4^ |
| *ABCC8* | F1182L, c.3546C>A | Reported in ^4^ |
| *ABCC8* | R1183W, c.3547C>T | Reported in ^36^ |
| *ABCC8* | R1183Q, c.3548G>A | Reported in ^38^ |
| *ABCC8* | P1199S, c.3595C>T | Reported in ^4^ |
| *ABCC8* | P1199Q, c.3596C>A | Reported in ^4^ |
| *ABCC8* | P1199L, c.3596C>T | Reported in ^47^ |
| *ABCC8* | G1256S, c.3766G>A | Reported in ^48^ |
| *ABCC8* | L1295F, c.3883C>T | Reported in ^4^ |
| *ABCC8* | R1314H, c.3941G>A | Reported in ^36^ |
| *ABCC8* | R1380C, c.4138C>T | Reported in ^38^ |
| *ABCC8* | R1380H, c.4139G>A | Reported in ^38^ |
| *ABCC8* | R1380P, c.4139G>C | Reported in ^4^ |
| *ABCC8* | R1380L, c.4139G>T | Reported in ^36^ |
| *ABCC8* | T1381N, c.4142C>A | Reported in ^4^ |
| *ABCC8* | S1501R, c.4503C>A | Reported in ^49^ |
| *ABCC8* | E1507G, c.4520A>G | Reported in ^50^ |
| *ABCC8* | E1507D, c.4521G>T | Reported in ^50^ |
| *ABCC8* | V1523M, c.4567G>A | Reported in ^51^ |
| *ABCC8* | V1524M, c.4570G>A | Reported in ^52^ |
| *ABCC8* | A1537P, c.4609G>C | Reported in ^53^ |
| *ABCC8* | V1540M, c.4618G>A | Reported in ^4^ |
| *INS* | ?, c.188-31G>A | Reported in ^54^ |
| *INS* | A24D, c.71C>A | Reported in ^55^ |
| *INS* | A24V, c.71C>T | Reported in ^56^ |
| *INS* | L30Q, c.89T>A | Reported in ^57^ |
| *INS* | G32S, c.94G>A | Reported in ^55^ |
| *INS* | G32R, c.94G>C | Reported in ^55^ |
| *INS* | C43S, c.127T>A | Reported in ^58^ |
| *INS* | C43G, c.127T>G | Reported in ^55^ |
| *INS* | F48C, c.143T>G | Reported in ^59^ |
| *INS* | R89C, c.265C>T | Reported in ^55^ |
| *INS* | C96Y, c.287G>A | Reported in ^55^ |
| *INS* | C96S, c.287G>C | Reported in ^59^ |
| *INS* | C96R, c.286T>C | Reported in ^58^ |
| *KCNJ11* | L17P, c.50T>C | Reported in ^4^ |
| *KCNJ11* | Q30_R34del, c.81_95del | Reported in ^60^ |
| *KCNJ11* | K39R, c.116A>G | Reported in ^43^ |
| *KCNJ11* | C42R, c.124T>C | Reported in ^33^ |
| *KCNJ11* | H46Y, c.136C>T | Reported in ^61^ |
| *KCNJ11* | H46L, c.137A>T | Reported in ^62^ |
| *KCNJ11* | N48I, c.143A>T | Reported in ^63^ |
| *KCNJ11* | I49F, c.145A>T | Reported in ^4^ |
| *KCNJ11* | R50G, c.148C>G | Reported in ^46^ |
| *KCNJ11* | R50Q, c.149G>A | Reported in ^61^ |
| *KCNJ11* | R50P, c.149G>C | Reported in ^64^ |
| *KCNJ11* | E51A, c.152A>C | Reported in ^65^ |
| *KCNJ11* | E51G, c.152A>G | Reported in ^4^ |
| *KCNJ11* | Q52R, c.155A>G | Reported in ^66^ |
| *KCNJ11* | Q52L, c.155A>T | Reported in ^67^ |
| *KCNJ11* | G53S, c.157G>A | Reported in ^68^ |
| *KCNJ11* | G53R, c.157G>C | Reported in ^68^ |
| *KCNJ11* | G53D, c.158G>A | Reported in ^61^ |
| *KCNJ11* | G53V, c.158G>T | Reported in ^69^ |
| *KCNJ11* | V59M, c.175G>A | Reported in ^66^ |
| *KCNJ11* | V59A, c.176T>C | Reported in ^47^ |
| *KCNJ11* | V59G, c.176T>G | Reported in ^66^ |
| *KCNJ11* | F60Y, c.179T>A | Reported in ^70^ |
| *KCNJ11* | V64M, c.190G>A | Reported in ^47^ |
| *KCNJ11* | W68R, c.202T>C | Reported in ^71^ |
| *KCNJ11* | W68G, c.202T>G | Reported in ^72^ |
| *KCNJ11* | V129M, c.385G>A | Reported in ^4^ |
| *KCNJ11* | A161T, c.481G>A | Reported in ^73^ |
| *KCNJ11* | L164P, c.491T>C | Reported in ^61^ |
| *KCNJ11* | C166Y, c.497G>A | Reported in ^61^ |
| *KCNJ11* | C166F, c.497G>T | Reported in ^74^ |
| *KCNJ11* | I167L, c.499A>C | Reported in ^75^ |
| *KCNJ11* | M169T, c.506T>C | Reported in ^4^ |
| *KCNJ11* | K170T, c.509A>C | Reported in ^61^ |
| *KCNJ11* | K170R, c.509A>G | Reported in ^40^ |
| *KCNJ11* | K170N, c.510G>C | Reported in ^53^ |
| *KCNJ11* | A174G, c.521C>G | Reported in ^46^ |
| *KCNJ11* | E179K, c.535G>A | Reported in ^4^ |
| *KCNJ11* | E179A, c.536A>C | Reported in ^38^ |
| *KCNJ11* | I182V, c.544A>G | Reported in ^68^ |
| *KCNJ11* | I182T, c.545T>C | Reported in ^76^ |
| *KCNJ11* | K185Q, c.553A>C | Reported in ^77^ |
| *KCNJ11* | K185T, c.554A>C | Reported in ^73^ |
| *KCNJ11* | R201S, c.601C>A | Reported in ^78^ |
| *KCNJ11* | R201G, c.601C>G | Reported in ^47^ |
| *KCNJ11* | R201C, c.601C>T | Reported in ^66^ |
| *KCNJ11* | R201H, c.602G>A | Reported in ^66^ |
| *KCNJ11* | R201L, c.602G>T | Reported in ^79^ |
| *KCNJ11* | E227K, c.679G>A | Reported in ^80^ |
| *KCNJ11* | E229K, c.685G>A | Reported in ^38^ |
| *KCNJ11* | L233F, c.697C>T | Reported in ^81^ |
| *KCNJ11* | V252M, c.754G>A | Reported in ^65^ |
| *KCNJ11* | V252L, c.754G>C | Reported in ^82^ |
| *KCNJ11* | V252A, c.755T>C | Reported in ^83^ |
| *KCNJ11* | V252G, c.755T>G | Reported in ^65^ |
| *KCNJ11* | P254Q, c.761C>A | Reported in ^84^ |
| *KCNJ11* | I284F, c.850A>T | Reported in ^85^ |
| *KCNJ11* | E292G, c.875A>G | Reported in ^83^ |
| *KCNJ11* | T293N, c.878C>A | Reported in ^86^ |
| *KCNJ11* | I296L, c.886A>C | Reported in ^66^ |
| *KCNJ11* | E322K, c.964G>A | Reported in ^87^ |
| *KCNJ11* | V328M, c.982G>A | Reported in ^4^ |
| *KCNJ11* | Y330C, c.989A>G | Reported in ^88^ |
| *KCNJ11* | S331P, c.991T>C | Reported in ^4^ |
| *KCNJ11* | F333L, c.997T>C | Reported in ^89^ |
| *KCNJ11* | G334S, c.1000G>A | Reported in ^4^ |
| *KCNJ11* | G334R, c.1000G>C | Reported in ^4^ |
| *KCNJ11* | G334C, c.1000G>T | Reported in ^73^ |
| *KCNJ11* | G334D, c.1001G>A | Reported in ^90^ |
| *KCNJ11* | G334V, c.1001G>T | Reported in ^91^ |

**Supplementary Table 5**: Known disease-causing dominant variants called by GATK3 mutect2. The coding location of variants are provided according to the following transcripts *ABCC8*: NM_001287174.1, *GCK*: NM_000162.3, *GLUD1*: NM_005271.4. The genomic locations of the *HK1* variants are provided according to GRCh38. Data on the alternative (ALT) and reference (REF) allele counts, along with the variant allele frequencies (VAF) and 95% confidence intervals (CI) are provided for both the tNGS and ddPCR data. Rows which are not highlighted denote variants that were confirmed by ddPCR (true positives), whilst rows highlighted in grey represent false positive calls that were not confirmed by ddPCR. ^‡^Indicates the two variants identified in one individual. ^†^Indicates variants not called by GATK4 mutect2. ^§^Samples with evidence of evidence of low-level contamination. *Low ddPCR VAF likely due to binding of reference allele probe to alternate allele probe. Unable to control out. **One replicate due to failing of column A.

| **Disease** | **Gene** | **Variant** | **tNGS ALT/REF allele count** | **tNGS VAF % (95% CI)** | **ddPCR ALT/REF droplets** | **ddPCR % (95% CI)** |
| --- | --- | --- | --- | --- | --- | --- |
| HI | *ABCC8* | c.4454G>A, p.(Gly1485Glu) | 27/317 | 7.8 (5.2-11.2) | 637/5027 | 11.2 (10.4-12.1) |
| HI | *ABCC8* | c.4151G>A, p.(Gly1384Glu) | 70/940 | 6.9 (5.4-8.7) | 489/8156 | 5.7 (5.2-6.2) |
| HI | *GCK* | c.641A>G, p.(Tyr214Cys) | 28/401 | 6.5 (4.4-9.3) | 476/7890 | 5.7 (5.2-6.2) |
| HI | *HK1* | Chr10:g.69348891C>T | 31/471 | 6.2 (4.2-8.7) | 166/3261 | 4.8 (4.1-5.6) |
| HI | *GCK* | c.1361_1363dup, p.(Ala454dup) | 59/932 | 6.0 (4.6-7.6) | 352/5917 | 5.6 (5.1-6.2) |
| HI | *GLUD1* | c.965G>A, p.(Arg322His) | 80/1272 | 5.9 (4.7-7.3) | 340/15993 | 2.1 (1.9-2.3)* |
| HI | *GLUD1* | c.1493C>T, p.(Ser498Leu) | 18/303 | 5.6 (3.4-8.7) | 418/7620 | 5.2 (4.7-5.7) |
| HI | *GLUD1* | c.1493C>T, p.(Ser498Leu) | 44/870 | 4.8 (3.5-6.4) | 237/4471 | 5.0 (4.4-5.7) |
| HI | *HK1* | Chr10:g.69348891C>T | 29/630^†^ | 4.4 (3.0-6.3) | 40/5936 | 0.7 (0.5-0.9) |
| HI | *HK1* | Chr10:g.69348932_69348935del | 21/486 | 4.1 (2.6-6.3) | 292/4936 | 5.6 (5.0-6.2) |
| HI | *GLUD1* | c.820C>T, p.(Arg274Cys) | 22/514 | 4.1 (2.6-6.1) | 269/5011 | 5.1 (4.5-5.7) |
| HI | *GLUD1* | c.953G>A, p.(Arg318Lys) | 21/494 | 4.1 (2.5-6.2) | 317/6277 | 4.8 (4.3-5.4) |
| HI | *GLUD1* | c.1466C>T, p.(Pro489Leu) | 15/386^†^ | 3.7 (2.1-6.1) | 343/9759 | 3.4 (3.1-3.8) |
| HI | *GLUD1* | c.1493C>T, p.(Ser498Leu) | 9/252^†^ | 3.4 (1.6-6.4) | 783/14632 | 5.1 (4.7-5.4) |
| HI | *GCK* | c.1361_1363dup, p.(Ala454dup) | 9/287 | 3.0 (1.4-5.6) | 545/12636 | 4.1 (3.8-4.5) |
| HI | *HK1* | Chr10:g.69348932_69348935del^‡^ | 13/432 | 2.9 (1.6-4.9) | 204/4999 | 3.9 (3.4-4.5) |
| HI | *GLUD1* | c.1519C>T, p.(His507Tyr) | 18/693 | 2.5 (1.5-4.0) | 177/5533 | 3.1 (2.7-3.6) |
| HI | *GLUD1* | c.1498G>A, p.(Ala500Thr) | 18/724 | 2.4 (1.4-3.8) | 157/6899 | 2.2 (1.9-2.6) |
| HI | *GCK* | c.1361_1363dup, p.(Ala454dup) | 9/512 | 1.7 (0.8-3.3) | 187/9965 | 1.8 (1.6-2.1) |
| HI | *GLUD1* | c.943C>T, p.(His315Tyr) | 12/795^†^ | 1.5 (0.8-2.6) | 88/17944 | 0.5 (0.4-0.6)* |
| HI | *GLUD1* | c.965G>A, p.(Arg322His) | 20/1517^†^ | 1.3 (0.8-2.0) | 52/6759 | 0.8 (0.6-1.0)* |
| HI | *GLUD1* | c.1493C>T, p.(Ser498Leu) | 4/306^†^ | 1.3 (0.4-3.3) | 344/15474 | 2.2 (2.0-2.4) |
| HI | *HK1* | Chr10:g.69348932_69348935del | 8/624 | 1.3 (0.5-2.5) | 309/9588 | 3.1 (2.8-3.5) |
| HI | *GCK* | c.1340G>T, p.(Arg447Leu) | 7/599^†^ | 1.2 (0.5-2.4) | 15/2626** | 0.6 (0.3-0.9) |
| HI | *ABCC8* | c.2143G>A, p.(Val715Met)^‡^ | 4/343 | 1.2 (0.3-2.9) | 1/4842 | 0.0 (0.0-0.1) |
| HI | *GLUD1* | c.1493C>T, p.(Ser498Leu) | 4/356^†^ | 1.1 (0.3-2.8) | 182/13198 | 1.4 (1.2-1.6) |
| HI | *ABCC8* | c.4433G>T, p.(Gly1478Val) | 7/628^†§^ | 1.1 (0.4-2.3) | 0/11232 | 0.0 (0.0-0.0) |
| HI | *GLUD1* | c.965G>A, p.(Arg322His) | 8/745 | 1.1 (0.5-2.1) | 46/9614 | 0.5 (0.3-0.6)* |
| HI | *ABCC8* | c.1433C>A, p.(Ala478Asp) | 6/676^†§^ | 0.9 (0.3-1.9) | 0/31334 | 0.0 (0.0-0.0) |
| HI | *ABCC8* | c.2143G>A, p.(Val715Met) | 4/464 | 0.9 (0.2-2.2) | 0/8749 | 0.0 (0.0-0.0) |
| HI | *ABCC8* | c.1432G>A, p.(Ala478Thr) | 4/531^†^ | 0.7 (0.2-1.9) | 0/10653 | 0.0 (0.0-0.0) |
| HI | *ABCC8* | c.4151G>A, p.(Gly1384Glu) | 4/554^†^ | 0.7 (0.2-1.8) | 1/9337 | 0.0 (0.0-0.1) |
| HI | *ABCC8* | c.4435G>A, p.(Gly1479Arg) | 4/565^†^ | 0.7 (0.2-1.8) | 1/9831 | 0.0 (0.0-0.1) |
| HI | *GCK* | c.194C>T, p.(Thr65Ile) | 3/525^†^ | 0.6 (0.1-1.7) | 1/9500 | 0.0 (0.0-0.1) |
| HI | *ABCC8* | c.2147G>A, p.(Gly716Asp) | 6/607^†^ | 0.5 (0.1-1.4) | 1/7382 | 0.0 (0.0-0.1) |
| **ddPCR not possible** | | | | | | |
| HI | *GLUD1* | c.943C>T, p.(His315Tyr) | 8/688^†^ | 1.2 (0.5-2.3) | Not performed, insufficient DNA | |
| HI | *GLUD1* | c.1519C>T, p.(His507Tyr) | 9/827^†^ | 1.1 (0.5-2.0) | Not performed, insufficient DNA | |
| HI | *GCK* | c.205T>C, p.(Ser69Pro) | 4/510^†^ | 0.8 (0.2-2.0) | Not performed, assay failed | |
| HI | *ABCC8* | c.4519G>A, p.(Glu1507Lys) | 3/410^†^ | 0.7 (0.2-2.1 | Not performed, insufficient DNA | |
| HI | *ABCC8* | c.4463A>G, p.(Gly1488Arg) | 3/432^†^ | 0.7 (0.1-2.0) | Not performed, insufficient DNA | |

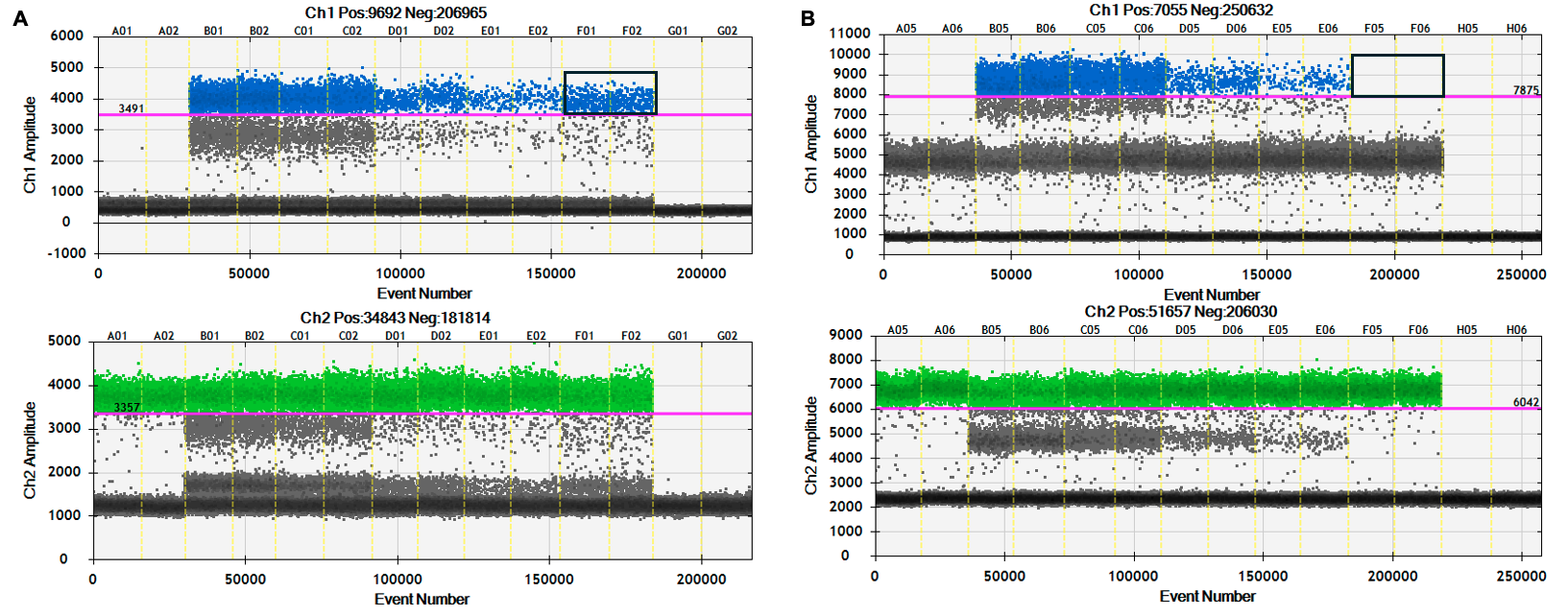

**Supplementary Figure 1**: Example droplet digital PCR (ddPCR) results. Each sample was tested in duplicate. Left to right: normal control without variant of interest (columns A), sample from case heterozygous for variant of interest (B), dilution of heterozygous control to create 25% control (C), 5% control (D), 2% control (E), case to test for the variant of interest (F), and NTC (water) (G). The blue droplets indicate the alternate allele and the green the reference allele. The purple line is the threshold, set at a level in which the 50%, 25%, 5% and 2% controls were close to those values, excluding the ddPCR ‘rain’. A) Example of a positive ddPCR result. Black box indicates droplets positive for alternate allele in the patient sample. B) Example of a negative ddPCR result. Black box indicates lack of droplets positive for alternate allele in the patient sample.

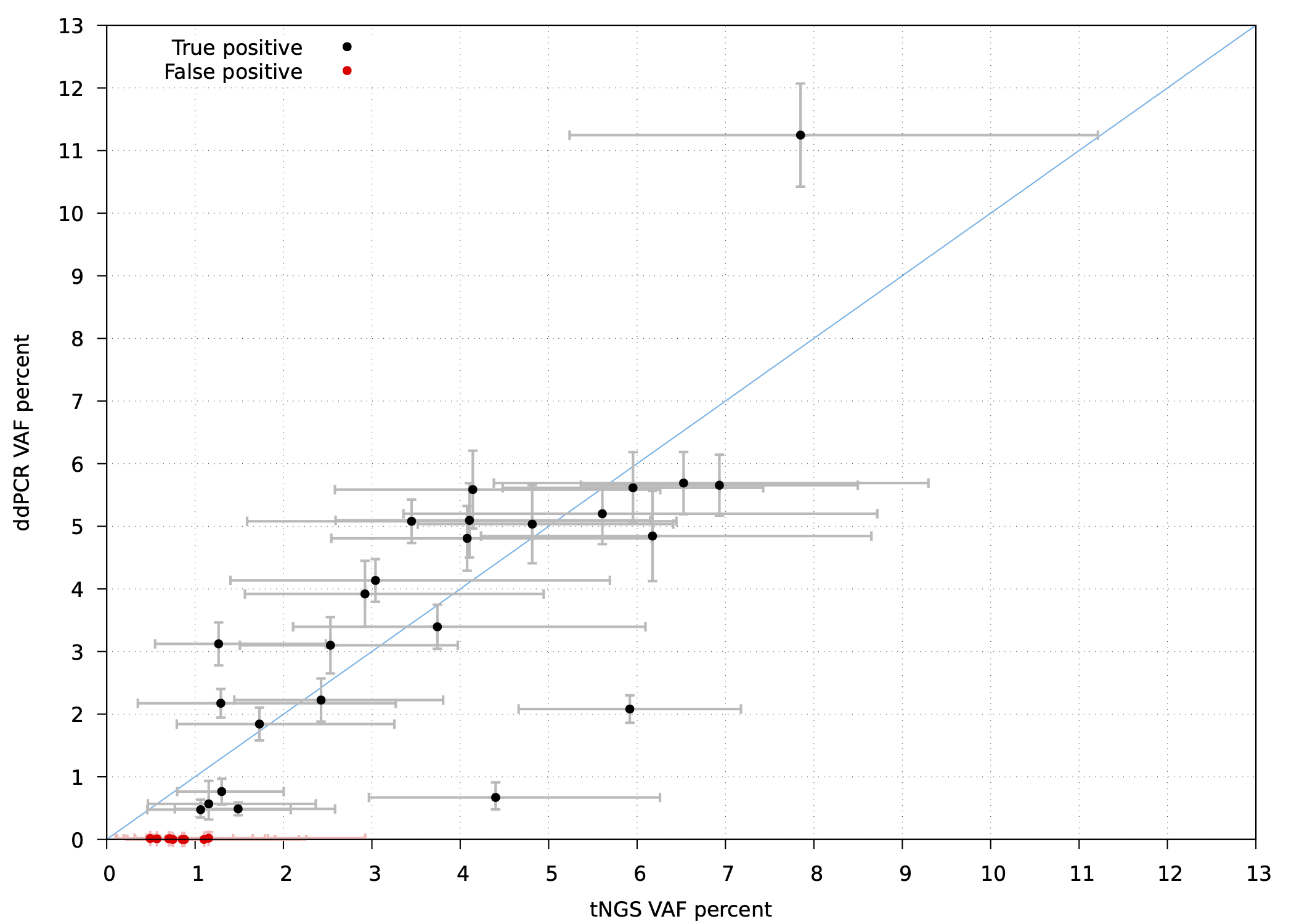

**Supplementary Figure 2**: Scatter plot of targeted next generation sequencing variant allele frequency (VAF) verses droplet digital PCR VAF. Error bars indicate 95% confidence intervals. Black: true positive variants confirmed by droplet digital PCR (ddPCR). Red: false positive variants not confirmed by ddPCR. R-squared value 0.6. Variation in concordance between tNGS and ddPCR can be explained by technical reasons such as sequence context impacting tNGS variant calls e.g. 4bp *HK1* deletion, and low ddPCR VAF compared to tNGS VAF in some samples due to binding of reference allele probe to alternate allele probe.

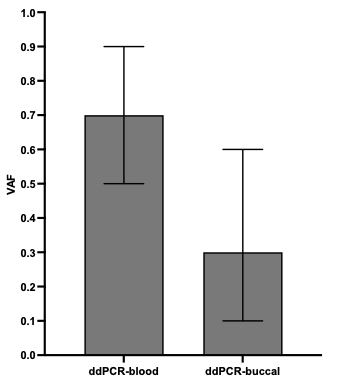

**Supplementary Figure 3:** ddPCR variant allele frequency (VAF) detected from blood and buccal DNA from one individual with a low-level mosaic *HK1* variant (Chr10:g.69348891C>T) Error bars indicate 95% confidence intervals.

**The International Congenital Hyperinsulinism Consortium**

Teoman Akçay^1^, Doha Sager Alhomaidah^2^, Nisha Bhavani^3^, Bianca Fiorella Miranda Cabrera^4^, Korcan Demir^5^, Ghaisana Fadiana^6^, Ghadir Elias-Assad^7,8^, Eiroa Hernan^10^, Rajesh Joshi^11^, Manjiri Karlekar^12^, Natalya Karp^13,14^, Vjosa Mulliqi Kotori^15^, Veronica Mericq^16^, Chirantap Oza^17^, Vaman Khadilkar^17^, VP Praveen^3^, Birgit Rami-Merhar^18^, Sumudu Nimali Seneviratne^19^, Zeynep Şıklar^20^, Yardena Tenenbaum Rakover^21^

**Affiliations**

1. Department of Pediatrics, Istanbul Health and Technology University (ISTUN) Medical Faculty, Istanbul, Türkiye.
2. Pediatric Endocrinology, Al Farwaniyah Hospital, Kuwait
3. Department of Endocrinology, Amrita Institute of Medical Sciences, Kochi, India
4. Pediatric Endocrinology Unit, Hospital Nacional Edgardo Rebagliati Martins, Lima, Peru
5. Department of Paediatric Endocrinology, Dokuz Eylül University, Izmir, Türkiye.
6. Cipto Mangunkusumo National General Hospital, Jakarta, Indonesia
7. Pediatric Endocrine Unit, Saint Vincent de Paul Hospital, Nazareth, Israel.
8. Azrieli Faculty of Medicine, Bar-Ilan University, Israel.
9. Department of Endocrinology, The Hospital for Sick Children, Toronto, Canada
10. Hospital Juan P. Garrahan, Ciudad de Buenos Aires, Argentina
11. B J wadia Hospital for Children, Acharya Donde Marg, Parel, Mumbai, India
12. Department of Endocrinology and Metabolism, Seth G. S. Medical College and KEM Hospital, Mumbai, India.
13. Western University, Department of Pediatrics, Division of Medical Genetics, London, Canada.
14. London Health Sciences Centre, Medical Genetics Program, Canada.
15. UBT College, Prishtina, Kosova
16. Institute of Maternal and Child Research, University of Chile, Santiago, Chile.
17. Jehangir Hospital, Pune, India
18. Dept. of Pediatric and Adolescent Medicine, Comprehensive Center for Pediatrics, Medical University of Vienna, Austria
19. Faculty of Medicine, University of Colombo, Colombo, Sri Lanka
20. Ankara University School of Medicine, Department of Pediatric Endocrinology, Ankara, Türkiye.
21. Consulting Center, Clalit Health Services, Afula, Israel
